# Community perceptions and attitude toward sexuality of women with disabilities in Kibra, Nairobi

**DOI:** 10.64898/2026.02.23.26346931

**Authors:** Brezhnev Henry Otieno, Sahaya Selvam

## Abstract

Despite the existence of strong global and national human rights frameworks that support disability inclusion, women with disabilities in Kenya are still heavily discriminated against and stigmatised due to the negative perceptions of the community. This study examined community attitudes toward the sexuality of women with disabilities in Kibra Sub-County, Nairobi, and investigated demographic factors influencing these views. Using a quantitative cross-sectional design, a stratified multistage random sample of 420 adult residents was surveyed using a perception questionnaire and the Attitudes Toward Disabled Person (ATDP) tool. The findings show that a large number of respondents recognize that women with disabilities have sexual feelings, have ‘normal’ sexual organs, and are sexually active. Even though most demographic variables did not have a significant association with perceptions of sexual activity, religion was one variable that had a significant association with perceptions of sexual anatomy. Overall attitudes towards women and men based on the ATDP test were positive as evidenced by mean ATDP scores for women (118. 76) and men (116. 36) which were above the respective standard thresholds (110 and 113). Multiple linear regression identified religion and education as significant negative predictors of positive attitudes, whereas close contact with persons with disabilities predicted more positive views. Their results indicated that the Kibra community, to some extent, recognizes the sexual agency of women with disabilities, nevertheless, this recognition is largely symbolic. In order to protect the sexual and reproductive rights of these women, the focus of the intervention should be shifted from the mere symbolic acceptance to the implementation of rights, based policies and culturally responsive strategies for inclusion in informal settlements.

## Introduction

The Convention on the Rights of Persons with Disabilities (CRPD), introduced a fresh perspective about disability across the globe. The CRPD established disability as a human rights matter instead of a medical classification or charitable treatment because it focuses on dignity, equality, and justice (1,2). Hence, people with disabilities are recognized as right holders and are therefore granted the fullest rights possible before the law. For women with disabilities, however, the realization of these rights is often restricted due to the intersection of gendered discrimination with ableist social norms. This particular intersection of exclusions is very visible in regard to sexual and reproductive health and rights (3,4).

The CRPD guarantees a huge array of protections, but in our context, Articles 23 and 25 become most pertinent as they underscore the right to family life and health services for women with disabilities (5). However, implementation has been uneven. Kenyan policies reflect the principles of the CRPD. However, the enforcement of sexual and reproductive health rights of women with disabilities is still limited. Negative attitudes from the public are common. Women with disabilities face a lot of stigma in health care settings. This poses even greater hindrances for them in trying to access basic services, including contraception and maternal care (6,7).

The contradictions between the public declarations in laws and realities on land indicate deeper structural imbalances. In most Kenyan society, cultural narratives link disability to supernatural causation, such as ancestor disobedience or witchcraft. These beliefs lead to the stigmatization of women with disabilities, who are often seen as unfit for motherhood or as threats to the well-being of others (8,9). Healthcare providers may deny them birth support, and families may isolate them out of fear that their impairment could affect individuals with disabilities (10).

Another difficulty is in the near absence of the hard-core empirical evidence on how CRPD obligations are being realized. Many reviews find very little clear evidence of successful interventions in settings to meet CRPD obligations (11,12). This lack of evidence would limit the formation of good policy and even weaken enforcement measures. This study attempts to add to the body of evidence on the sexual reproductive health and rights of women with disabilities in view of the provisions of the CRPD.

### Perspectives on disability and sexuality of women with disabilities Global context

Global research shows that women with disabilities consistently face barriers to sexual and reproductive healthcare due to the combined effects of ableism, gender inequality, and economic disadvantage.

Structural neglect in the healthcare systems is a widespread problem. Basic gynaecological tools are often missing in clinics, venues for treatment are hardly accessible, and providers are poorly trained with regard to disability-inclusive care (13). So many of them are not able to handle the concerns of women with disabilities. The other problem is clinical gatekeeping that deems women with disabilities as unfit to be mothers and restricts their reproductive choices (14). Such practices stem from cultural beliefs that either dismiss or problematize their sexuality, thus stripping the woman with disabilities of any autonomy or dignity.

Women with disabilities have been stereotyped again and again as asexual, not attractive, or non-feminine, and by such perceptions, these women are denied the right to motherhood and sexual agency (15). Such descriptions of women with disabilities become socially and institutionally recognized, furthering stigma against them and undermining their autonomy.

Healthcare environments themselves are steeped in ableism. Most caregivers are not appropriately trained and equipped to take care of and cater to the unique needs of women with disabilities. In addition to the physical impediments, much difficulty arises due to the perplexing insurance policies and the numerous bureaucratic layers obstructing the provision of care. Ableism also intersects with poverty, marital status, and power relations, generating compounded exclusion, especially in reproductive autonomy (16). This intersectionality means marginalization operates on multiple, overlapping levels.

The whole premise of sexual citizenship underscores the universality of sexual and reproductive rights (17). However, many women with disabilities are stripped of these very rights. Forced sterilizations, substituted decision-making, and medical paternalism are still operating under the thinly mitigated excuse of protection while really aiming at control (18,19). These measures are representative of the new forms of eugenic thinking. Women with disabilities are deemed incompetent to reproduce or to consent to matters related to their bodies, and on account of this, surveillance is imposed on them (18)

Controlling disability, The reproductive lives of women are an aspect of structural violence. The issue requires continuous attention to laws, policies, and institutional practices that continue to suppress their rights.

### Regional Context

In sub-Saharan Africa, women with disabilities encounter barriers impacting their lives, choices, bodies, and rights. Their experiences are largely determined by intersecting factors such as societal attitudes, perceptions, stigma, and, on occasions, the nature of their disabilities. These barriers extremely limit their possibilities to access sexual and reproductive health and rights.

One of the most typical barriers is the existence of cultural perceptions that desexualize women with disabilities. In rural Ghana, for instance, disability is conceived either spiritually or morally as punishment or bad fortune (8). These kinds of attitudes characterize the reproductive lives of disabled women as illegitimate or unsuitable. Pregnant women with disabilities are thus stereotyped as unclean and shunned by their families and society in general. These social exclusions not only delegitimize and disregard their sexuality and desire for motherhood but also perpetuate emotional isolation.

The exclusion and marginalization of women with disabilities is made worse by unresponsive health systems. Some of the facilities are physically inaccessible and lack ramps or sign language interpretation (20,21). Attitudes of healthcare providers create barriers for individuals. Some providers hold moralistic or paternalistic views that prohibit open discussions about sexual health, consent, and pleasure (22,23).

These institutional barriers do not exist by themselves, as they stand on broader social and economic inequalities. The findings from Ghana illustrate how mothers with blindness in Ghana confront interlocking discrimination (24). These intersecting dimensions of disability, poverty, marital status, and dependence on others generate further exclusion for women with disabilities and limit their access to care.

Even when inclusive health policies exist, they are often poorly put into practice. In Uganda, for example, poor infrastructure, discrimination by service providers, and inadequate communication systems are chronic breakers of official promises of affordable health care (25). The findings from Southern Africa Thabethe indicate that women with disabilities are barely consulted in health decision-making. As a result, their needs are overlooked, and they are concealed in the very mechanisms meant to cater to them (26).

### Kenyan Context

The sexual and reproductive health and rights (SRHR) of Kenyan women with disabilities remain under-researched. The little academic literature that does exist documents marginalization of women with disabilities. They are habitually infantilized, their sexual agency is negated, and their reproductive health needs are ignored at societal and institutional levels.

Empirical studies reveal that exclusion from opportunities for women with disabilities is complex and multilayered. For example, in Kajiado County, it is observable that over 60% of women with disabilities held negative attitudes toward family planning (27). This resistance was not inherent, but it was borne of the stigma that society places upon the sexuality of people with disabilities and the use of contraception. They further observe that Only 32% of women and girls with disabilities used a contraceptive, which reflects dire failures in outreach, access, and trust in health systems. These findings place national health programs under scrutiny for inadequately responding to the sexual reproductive health and rights of women with disabilities. Recent research in Kenya indicate that women with disabilities are at times in search of sexual and reproductive health information while demanding their rights concerning all sexual matters. However, several structural barriers block them, such as familial resistance, poor infrastructural access, economic insecurity, and unfading social stigma (28). This is not just incidental but structural, shaped by dominating patriarchal and ableist ideologies. This denial of sexual and reproductive health and sexual rights for women with disabilities is not simply because of personal biases or neglect of institutions; it signifies deeper power structures that render their bodies marginal and deny them sexual autonomy.

Cultural discourses hold a central place in reproductive exclusion. It has been observed that disability in Kenya is often conceptualized through multiple cultural frameworks (29). When disabled persons tell their own stories publicly, the dominant story is lifted from medicalized or tragic interpretations. This kind of narrative promotes a broad and inclusive idea of personhood. Such testimonies, the authors claim, represent a political act that rejects deficit-based conceptualizations. They affirm the sexual agency of persons with disabilities.

Technological interventions such as assistive devices add complexity to how disability is experienced in social contexts. Prosthetic devices for lower-limb amputation are intended to restore mobility and support functional independence for users (30). However, research in Kenya has shown that use of assistive technologies including prostheses and mobility aids can attract stigma and unwanted attention in public spaces, leading some users to avoid social engagement or public environments (31). Therefore, while these devices support mobility and greater independence, they also operate as visible social markers that shape experiences of belonging and exclusion.

In religious contexts, faith traditions can support inclusion while also reinforcing exclusion of people with disabilities. In Western Kenya, some Christian interpretations link disability to moral failure or divine punishment, which contributes to discrimination and limits full participation in religious life and leadership for persons with disabilities (32). Grounded theology rooted in the lived experiences of people with disabilities has the potential to promote more inclusive participation in church life. From a regional perspective, religious communities often provide significant emotional and material support to individuals with disabilities (33). At the same time, certain segments of these communities perpetuate moral judgments that generate shame for persons with disabilities and deepen social exclusion. These two sets of findings highlight the dual role of religion: religious institutions can serve as sources of support while simultaneously contributing to marginalization.

Cumulatively, the Kenyan literature reveals intersecting institutional, cultural, and symbolic mechanisms that marginalize, desexualize, and dehumanize women with disabilities. However, none of them have used standardized measures such as the ATDP scale to evaluate attitudes toward individuals with disabilities. This study seeks to address this important gap.

The present study focuses on community perceptions and attitudes regarding the sexuality of women with disabilities in Kibera, an urban informal settlement in Nairobi. The study is organized around three primary objectives:

1. To establish community perception of the sexuality (sexual desires, sexual organs, and sexual intercourse) of women with disabilities.
2. To measure societal attitudes towards women with disabilities.
3. To investigate the sociodemographic factors associated with these attitudes and perceptions - sex, age, religion, and contact with individuals with disabilities.

## Methodology

### Study Design

A quantitative cross-sectional design was used to study community perceptions and attitudes toward women with disabilities. This design allowed data to be collected at a single point in time, providing efficiency and timeliness in assessing perceptions while enabling analysis of their relationships with demographic variables. This was the most practicable and fitting approach for addressing the research question (34,35).

### Study Setting

This research was conducted in Kibra Sub-County, Nairobi, one of the most densely inhabited informal settlements of Kenya (36). It was chosen because of its socio-economic diversity, high population density, and the obvious structural inequalities that mostly impact women and persons with disabilities. Informal urban settings such as Kibra tend to magnify social stigma, constrain service delivery options, and perpetuate stigma, hence becoming important scenario for disability research (37). In this context, Kibra was relevant for investigating community perceptions of sexuality of women with disabilities and attitudes towards them.

### Study Population and Sampling Strategy

The target population was Kibra adult residents and was picked from an estimated population of 185,777 (2019 Kenya Population and Housing Census). The sample size with a 95 percent confidence level and a 5 percent margin of error is computed below based on Yamane formula (38):

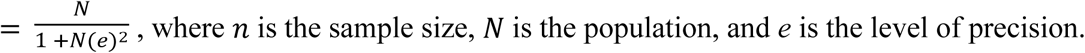

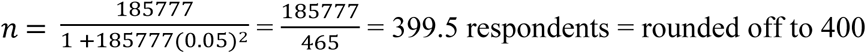

To adjust for anticipated non-response, particularly given the challenges typical in urban fieldwork and the sensitivity of the topic, the target sample size was increased by 45%, reaching 580 (39). A total of 420 respondents participated, yielding a response rate of 72.4%.

A multistage stratified random sampling approach was used. The sampling began with the identification of administrative units, followed by the random selection of enumeration areas within these units. Systematic sampling was applied for household selection. Stratification was done by gender to ensure proportional representation, according to local census details. Adult respondents without any disabilities were the only respondents selected for the final sample.

### Data Collection

The questionnaire contained demographic items, Likert-scale questions, and direct measures of perceptions relating to sexuality (sexual feelings, sexual organs, and sexual activities) of women with disabilities. The study utilized the Attitudes Toward Disabled Persons (ATDP) Form B (40), which was appropriately adapted to suit the local context, in order to assess general attitudes.

A short-term consultant was enlisted to oversee hourly data uploads to perform quality control in real-time. Depending on the choice of respondents, the questionnaires were administered in either the English or Swahili languages. The respondents voluntarily participated in the survey, and written informed consent was obtained from all. Anonymous unique identification codes were used to protect the identity of respondents, and no information that could be associated with a respondent’s identity was collected.

### Data Analysis

Data were analyzed using SPSS version 29. Descriptive statistics were used to summarize respondents’ demographic characteristics. Nonparametric methods, the Mann-Whitney U test and Kruskal-Wallis H test, were used to compare differences in ATDP mean scores across the demographic variables, given that the Shapiro-Wilk test rejected the null hypothesis of normality for ATDP scores (p < 0.01). The U test was applied for comparisons between two groups, whereas the H test was applied for comparisons among more than two groups. For Post hoc, the Dunn-Bonferroni test for pairwise comparisons was applied where the bivariate analyses were statistically significant.

Chi-square tests were applied to evaluate the link between categorical variables. Effect sizes were reported in terms of Phi (ϕ) and Cramer’s V (V). A multiple linear regression model was then run to identify independent predictors of ATDP scores, considering only those factors found significant in bivariate analyses.

### Ethical Considerations

The study was approved by Tangaza University and the National Commission for Science, Technology, and Innovation (NACOSTI; Permit No. 558428). Further permission was secured from the local administrative officers in Kibra. The participants were fully informed about the study’s goals and procedures. Each participant gave their written informed consent before participating in the research. Privacy and confidentiality were maintained throughout the data collection, while the data were anonymized right from the very beginning.

## Results

The findings are organized into four specific objectives. The first objective focused on perceptions of sexuality including, sexual feelings, sexual organs, and sexual activity. The second evaluated perception was on fertility and pregnancy. The third measured attitudes toward women with disabilities. The last investigated differences in perceptions and attitudes according to various sociodemographic characteristics. These are age, gender, educational level, religious affiliation, and nature of contact with people with disabilities

### Respondents Characteristics

A total of 420 out of the 580 sampled individuals completed the survey. The response rates for females and males were 71% and 74%, respectively. Most respondents were Christian (85.5%) and 10.4% were Muslim. A minority followed African Traditional Religion (2.9%), and a small proportion were not religious (1.2%.)

When asked about their interaction with persons with disabilities, just over half (51%) described their contact as casual. Nearly one-third (30%) reported being caregivers, while 18% stated they had no close contact. Levels of education were quite diverse; 39% had secondary education, 24.8% primary education, 19.5% some college, 9.3% had technical training, 25% had primary school, and 2% indicated not having any formal schooling. A detailed demographic distribution of the sample is shown in Table 1.

**Table 1:**
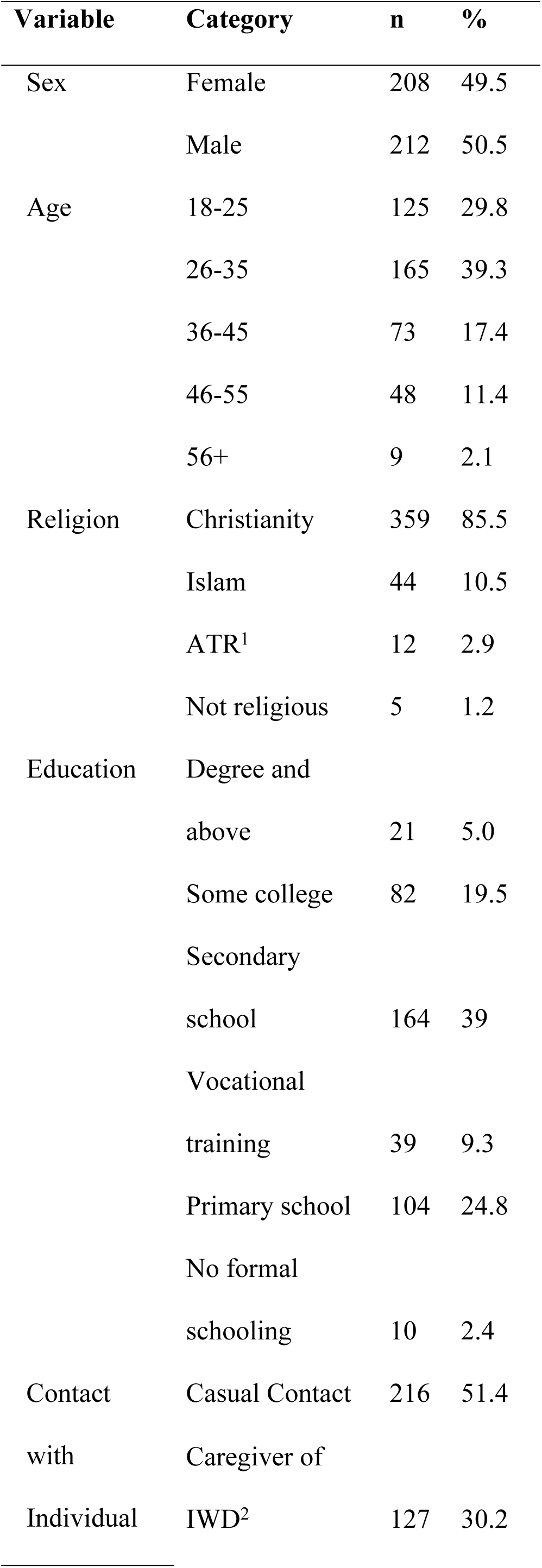
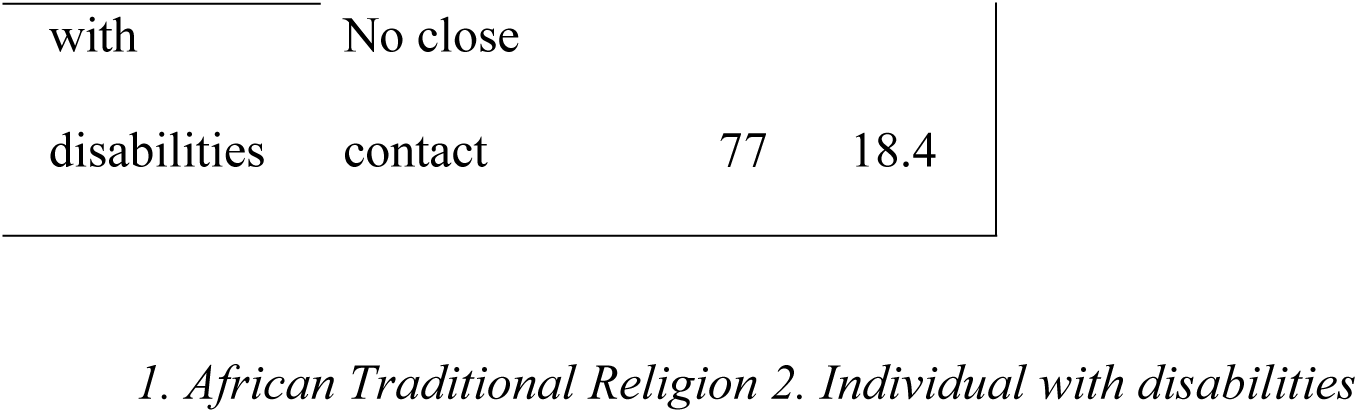
Background Characteristics of the Respondents (N = 420)

### Perceptions of Sexual Feelings

Pearson’s chi-square tests of independence were conducted to examine whether perceptions of sexual feelings in women with disabilities varied by demographic characteristics (N = 420). No statistically significant associations were found for sex, χ²(1) = 0.15, p = .695; age category, χ²(4) = 3.39, p = .495; religious affiliation, χ²(3) = 0.55, p = .907; educational background, χ²(5) = 3.51, p = .622; or contact with individuals with disabilities, χ²(2) = 3.02, p = .221.

The majority of respondents (95.2%) reported that women with disabilities have sexual feelings. The absence of statistically significant differences suggests that this perception is widely held and consistent across demographic groups, including age, sex, religion, education level, and contact with individuals with disabilities.

**Table 2:**
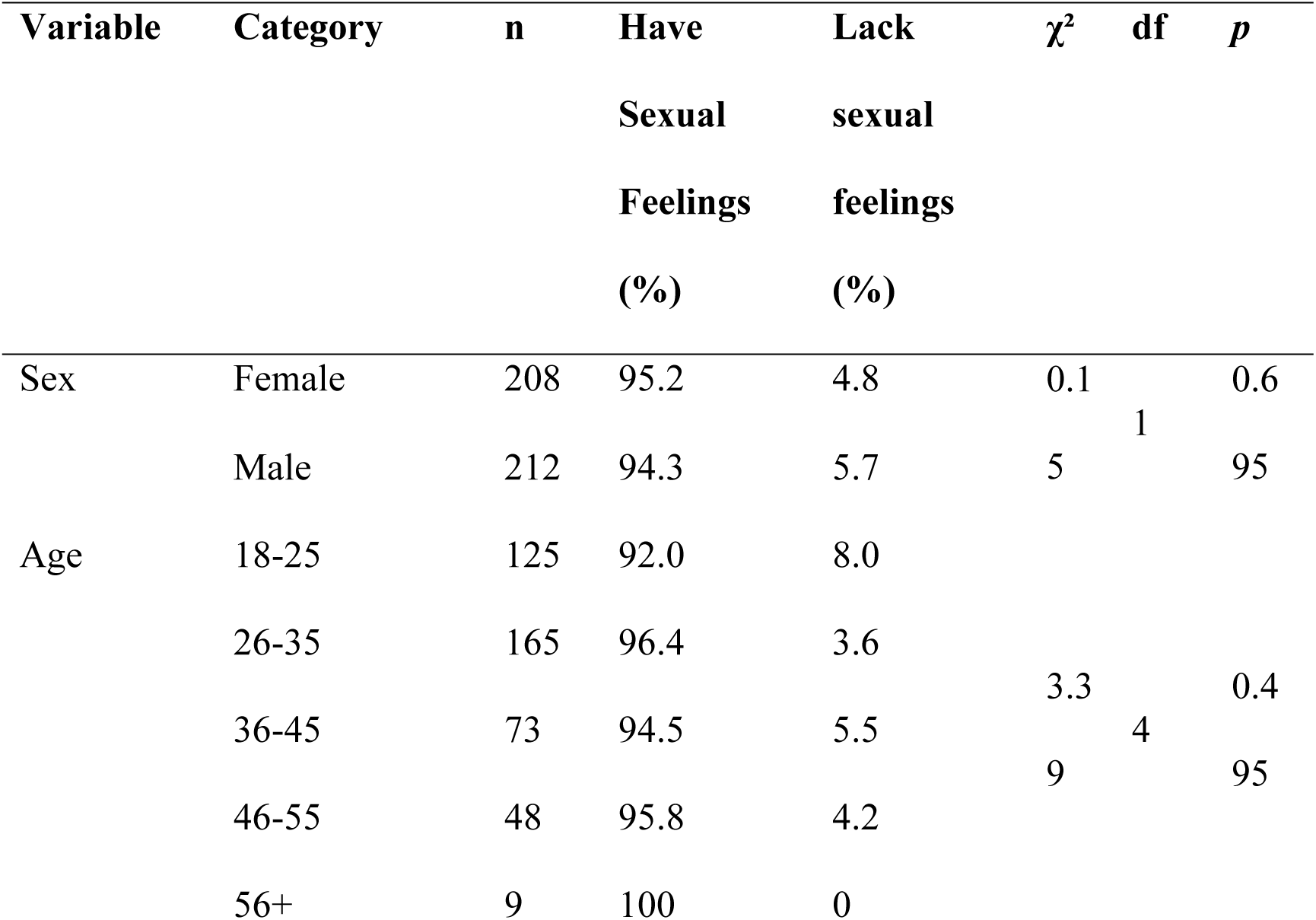
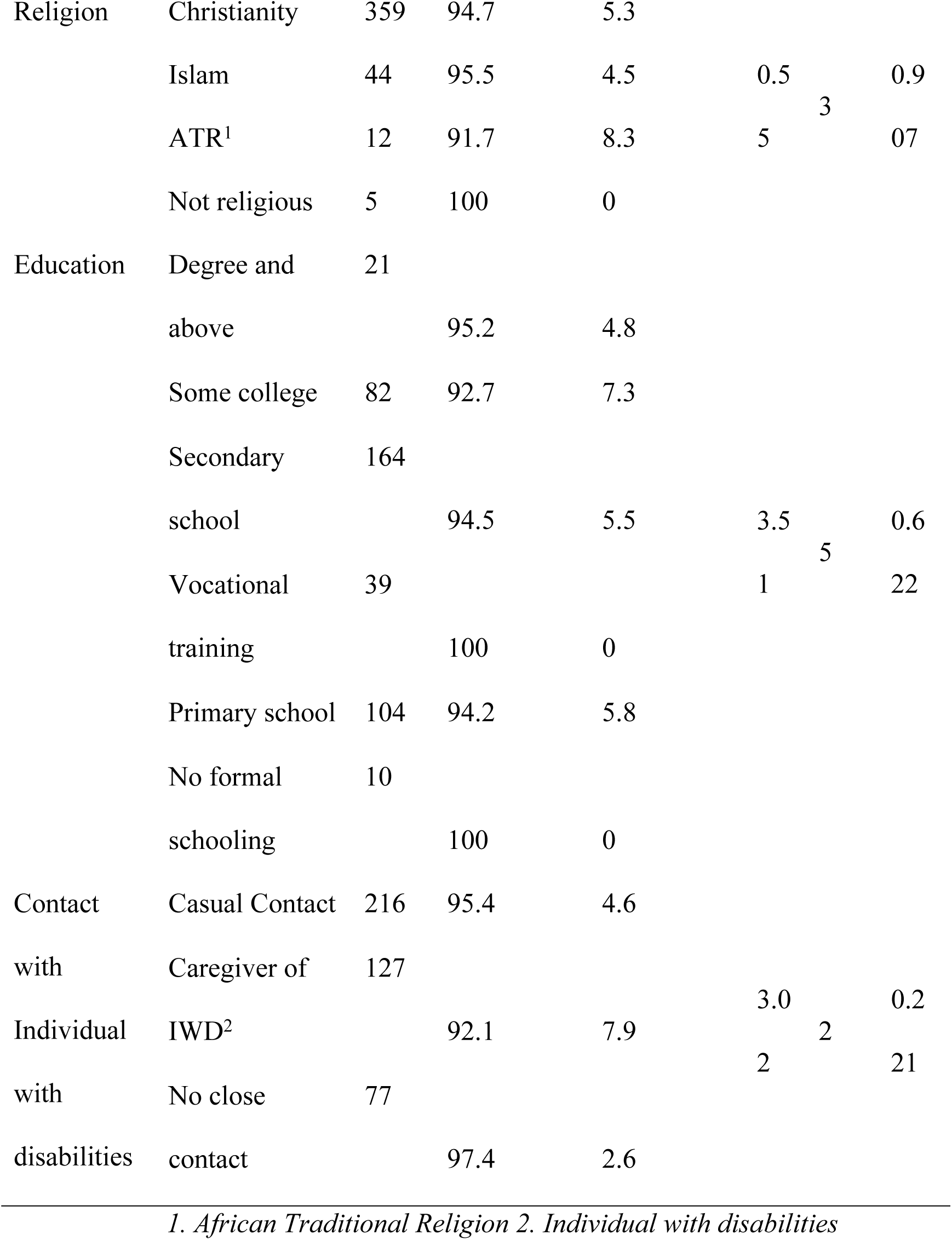
Demographic Characteristics and Perceptions of Sexual Feelings of Women With Disabilities (N = 420)

### Perceptions of Sexual organs

A series of Pearson’s chi-square tests of independence were conducted to examine whether demographic characteristics were associated with perceptions of sexual organs in women with disabilities (N = 420). A statistically significant association was observed between religion and perceptions of sexual anatomy, χ²(3) = 13.06, p = .005, with a small effect size (ϕ = 0.176). No significant associations were found for sex, χ²(1) = 0.72, p = .398; age category, χ²(4) = 8.92, p = .063; educational background, χ²(5) = 8.38, p = .137; or contact with individuals with disabilities, χ²(2) = 0.31, p = .985.

Nearly all participants (over 99%) perceived women with disabilities as having “normal” sexual organs. However, the significant influence of religious affiliation suggests that cultural or doctrinal beliefs may modestly shape how individuals evaluate sexual anatomy of women with disabilities. The lack of significant variation across other demographic factors indicates a largely homogeneous perception on this issue outside of religious differences.

**Table 3:**
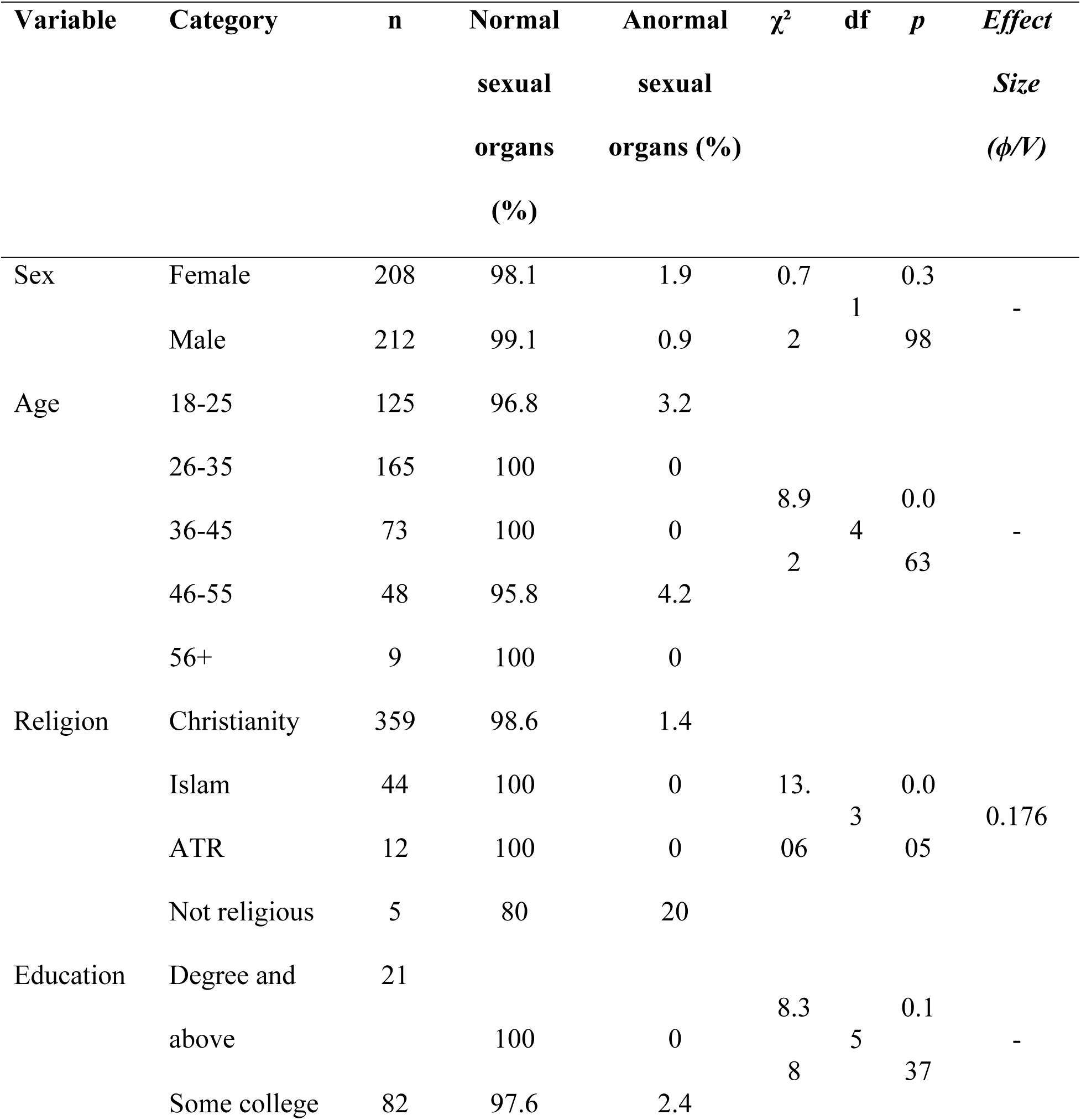
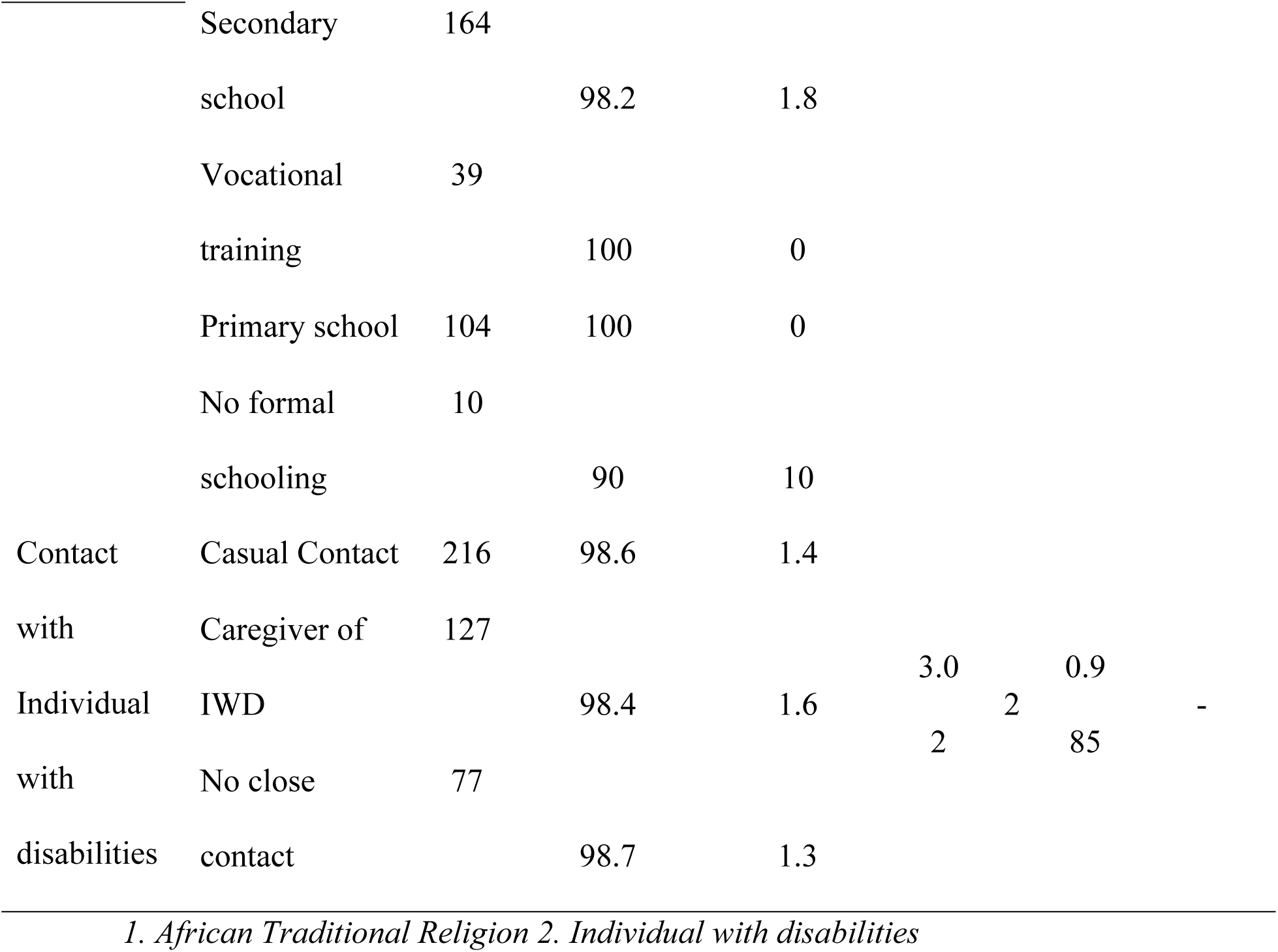
Demographic Characteristics and Perceptions of Sexual Organs of Women With Disabilities (N = 420)

### Perceptions of Sexual Activity

Pearson’s chi-square tests were conducted to assess whether demographic characteristics were associated with perceptions of sexual activity in women with disabilities (N = 420). The analyses revealed no statistically significant associations for sex, χ²(1) = 0.13, p = .722; age category, χ²(4) = 1.59, p = .811; religious affiliation, χ²(3) = 1.21, p = .751; educational background, χ²(5) = 1.93, p = .859; or contact with individuals with disabilities, χ²(2) = 1.05, p = .591.

The vast majority of respondents (97.6%) perceived women with disabilities as sexually active. The absence of significant variation across all demographic subgroups indicates a consistent and widely held belief in the sexual agency of women with disabilities. These findings suggest a normative recognition of sexual activity in this population, regardless of the respondent’s sex, age, religious orientation, educational attainment, or level of contact with individuals with disabilities.

**Table 4:**
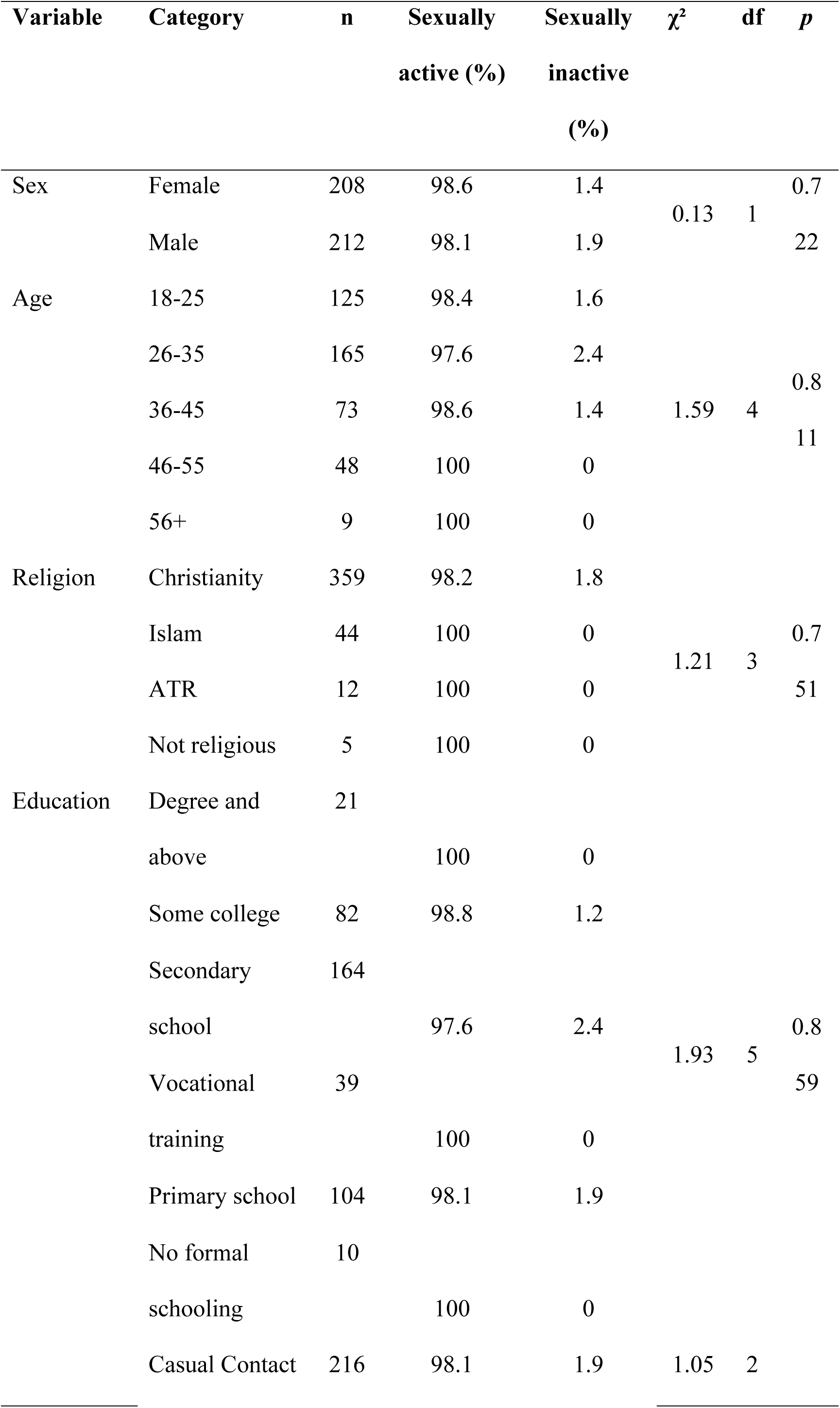
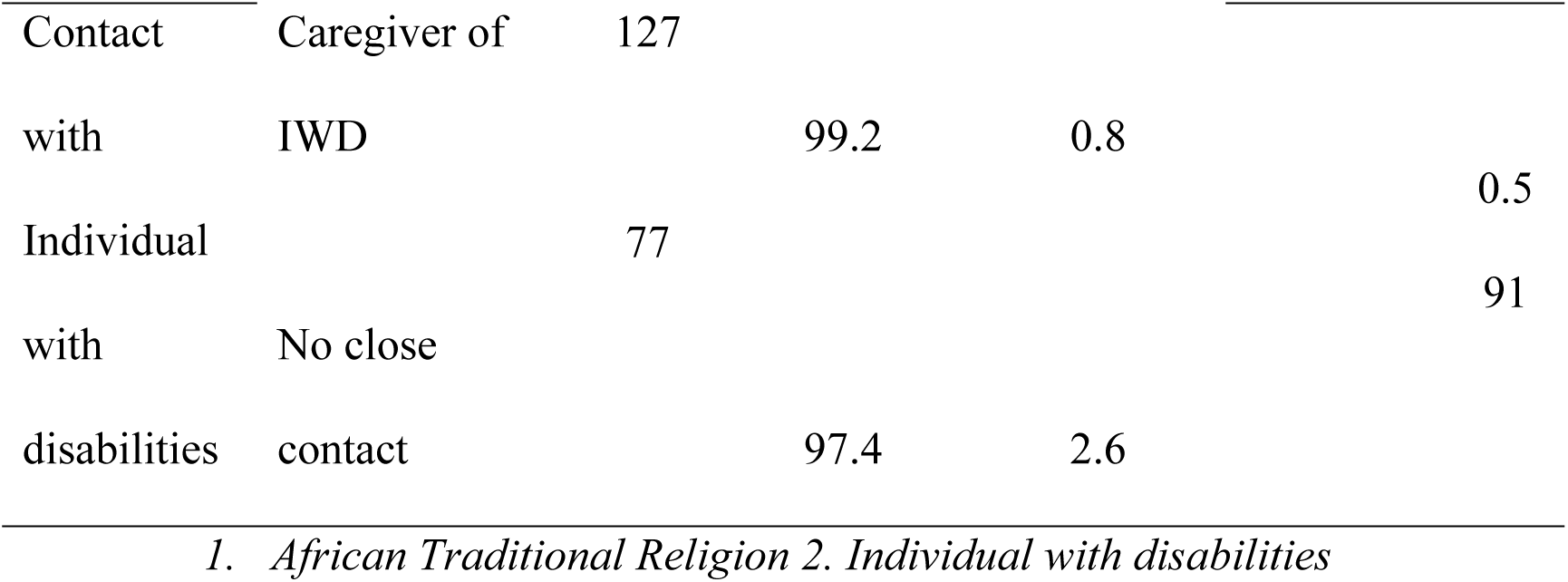
Demographic Characteristics and Perceptions of Sexual Activity of Women With Disabilities (N = 420)

### Attitudes Toward Women with Disabilities

#### Bivariate and Multivariate Analysis of ATDP Scores

The mean ATDP score across the sample was 117.55 (SD = 24.18), with a maximum observed score of 180 (maximum=, minimum=_. Female participants (M = 118.76, SD = 25.94) reported slightly higher scores than males (M = 116.36, SD = 22.39), although this difference was not statistically significant, U = 20592.50, p = .242. No significant differences were observed across age groups, H(4) = 1.68, p = .794.

However, significant variation in attitudes toward women with disabilities was found based on religion, H(3) = 16.83, p < .001; education, H(5) = 21.50, p < .001; and contact with individuals with disabilities, H(2) = 18.61, p < .001 (see Table 5).

**Table 5.**
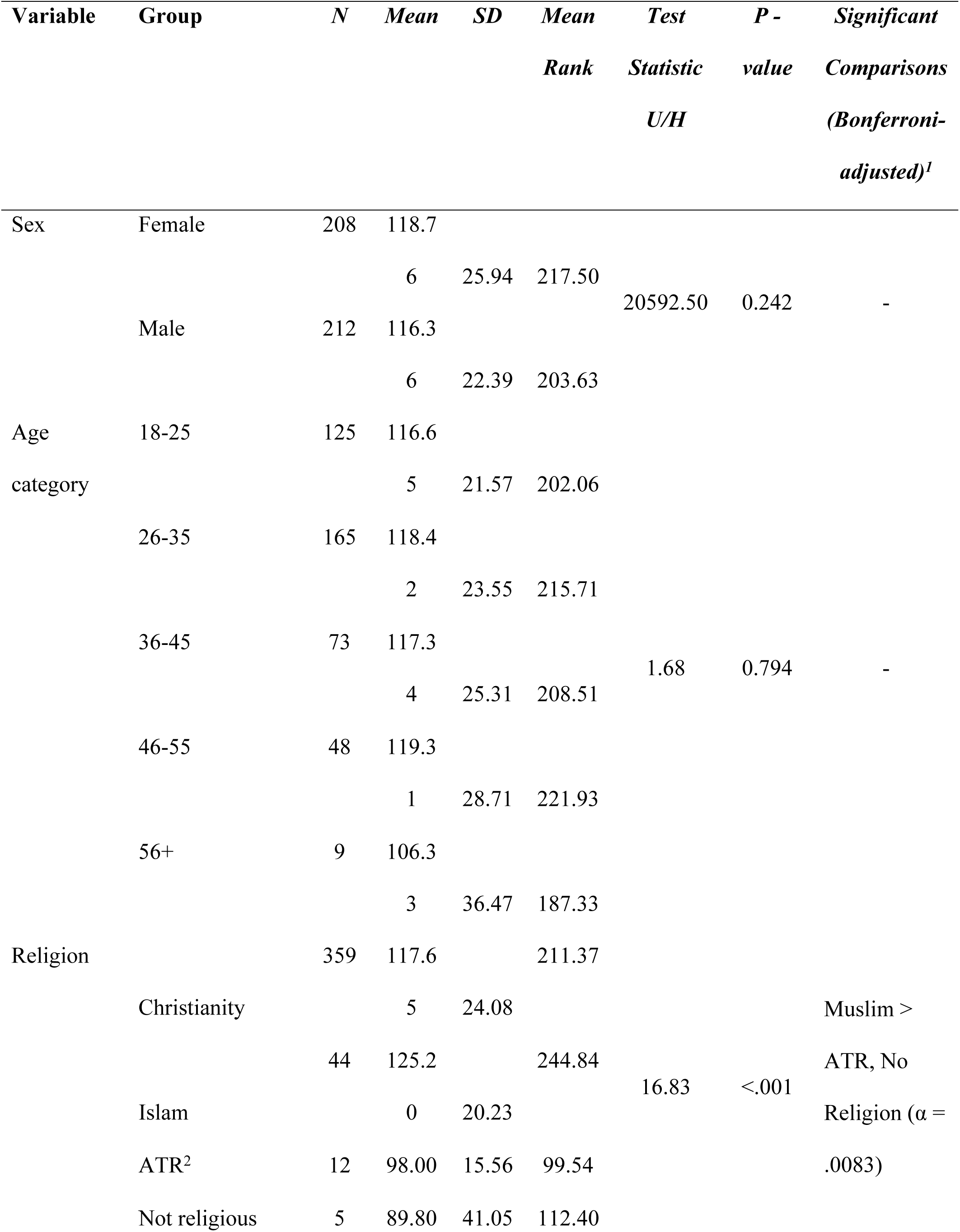
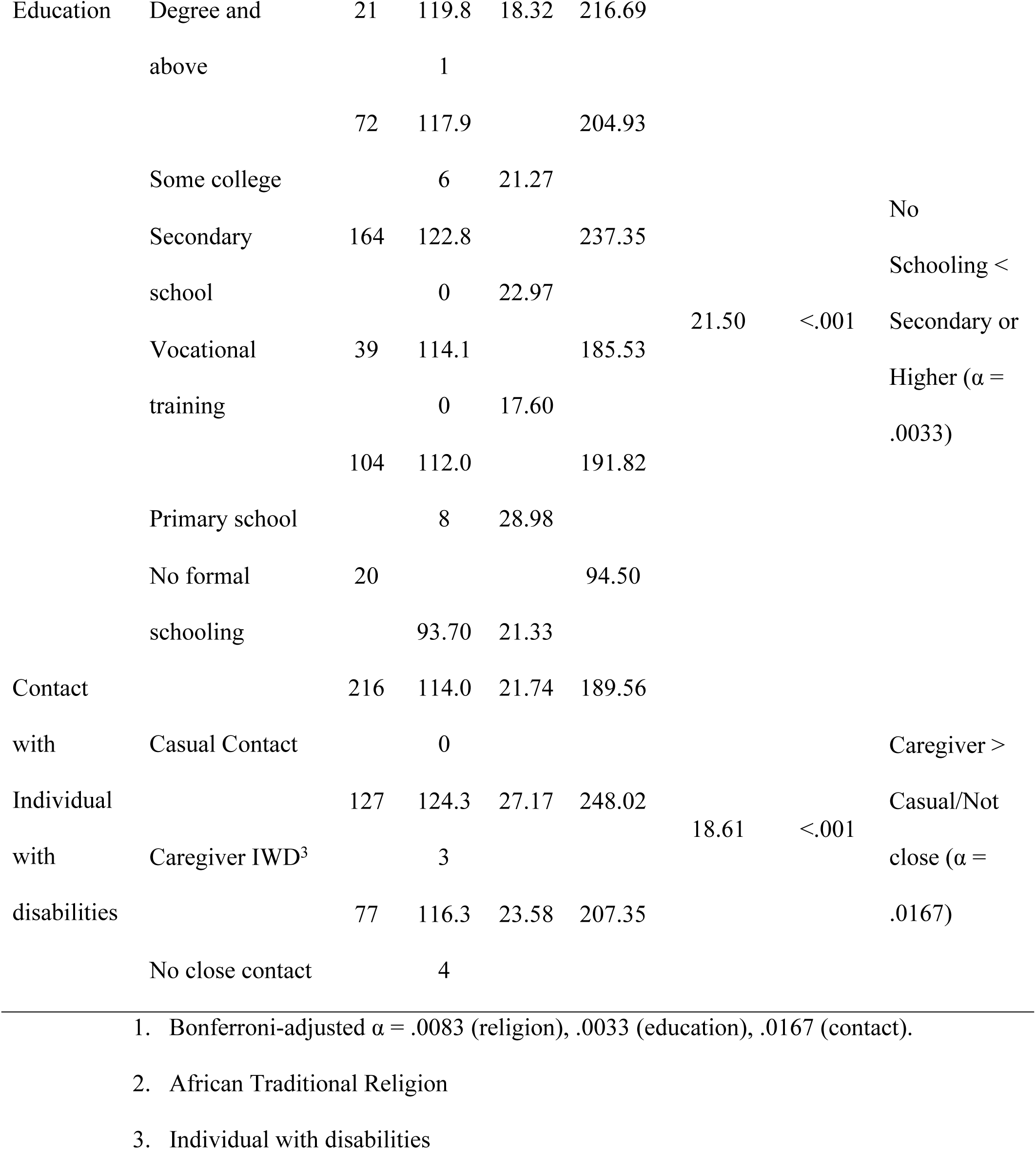
Bivariate Analysis of ATDP Scores by Demographic Variables. *Mann–Whitney U and Kruskal–Wallis Tests with Bonferroni-Adjusted Post Hoc Comparisons*

Bonferroni-adjusted post hoc comparisons indicated that:

- Muslim participants reported significantly more positive attitudes than those identifying with no religion (p_adj_ = .007) and African Traditional Religion (p_adj_ = .006).
- Participants with no formal schooling scored significantly lower than those with secondary (p_adj_ = .002) and tertiary education (p_adj_ = .001).
- Those with caregiving experience had significantly more favourable attitudes than individuals with casual (p_adj_ = .011) or no close contact (p_adj_ = .013).

To further examine these relationships, a multiple linear regression was conducted to predict ATDP scores using religion, education, and contact with individuals with disabilities as predictors. The model was statistically significant, F(3, 416) = 7.35, p < .001, and explained approximately 5% of the variance in ATDP scores (R² = .050, Adjusted R² = .044) (see Table 6).

**Table 6:** Model Summary: Multiple Linear Regression Predicting ATDP Scores. *Model Summary^b^*

| Model | R | R Square | Adjusted R Square | Std. Error of the Estimate |
| --- | --- | --- | --- | --- |
| 1 | .224<sup>a</sup> | 0.050 | 0.044 | 23.680 |

a. Predictors: (Constant), Contact with disabilities, Religious affiliation, Educational background
b. Dependent Variable ATDP Score

As shown in Table 7, religion (β = −.108, p = .024) and education (β = −.181, p = .001) were significant negative predictors of ATDP scores, indicating that certain religious and educational contexts may be associated with less favourable attitudes. In contrast, closeness of contact with individuals with disabilities emerged as a significant positive predictor (β = .107, p = .026), suggesting that more direct and personal interactions foster more positive attitudes.

**Table 7:**
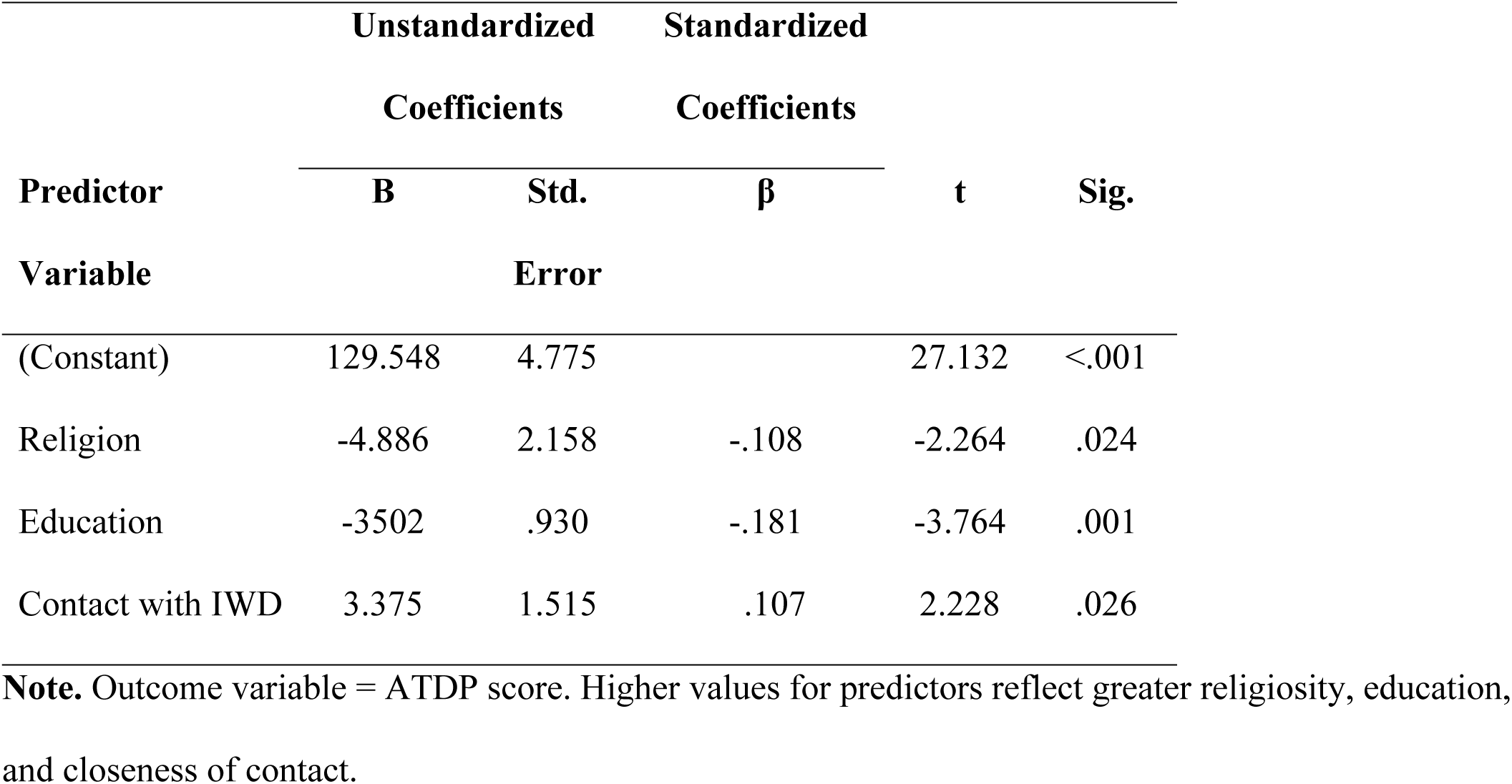
Regression Coefficients for Predictors of ATDP Scores. *Coefficients^a^*

| Predictor<br>Variable | Unstandardized<br>Coefficients |  | Standardized<br>Coefficients | t | Sig. |
| --- | --- | --- | --- | --- | --- |
| | B | Std. Error | $\beta$ | | |
| (Constant) | 129.548 | 4.775 |  | 27.132 | <.001 |
| Religion | -4.886 | 2.158 | -.108 | -2.264 | .024 |
| Education | -3502 | .930 | -.181 | -3.764 | .001 |
| Contact with IWD | 3.375 | 1.515 | .107 | 2.228 | .026 |

**Note.** Outcome variable = ATDP score. Higher values for predictors reflect greater religiosity, education, and closeness of contact.

Taken together, these results indicate that while some demographic factors (e.g., sex, age) are not significantly associated with attitudes toward persons with disabilities, while religious orientation, education, and type of contact with individual with disabilities demonstrate consistent and meaningful effects across both bivariate and multivariate models.

## Discussion

This study endeavoured to investigate the perceptions among community members in Kibra, Nairobi, on the sexuality, fertility, and pregnancy of women with disabilities. The study also measured the attitude of community members towards women with disabilities. The findings on the perceptions and the attitudes are discussed and compared with local, regional and global literature.

### Community perceptions of sexual feelings of women with disabilities

Community recognition in our study that women with disabilities experience sexual feelings marks a significant departure from long-standing narratives that have portrayed them as asexual. This shift likely reflects broader sociocultural changes. Recognition spans all demographic variables, sex, age, religion, education, and prior contact with disability. The findings point to growing awareness. But it remains important to ask, does this awareness translate into actual support for sexual autonomy and rights?

There is a growing body of evidence that shows how structural and normative barriers still continue to restrict access to sexual and reproductive health and rights for women with disabilities, Global studies as well as those from sub-Saharan Africa (including Kenya) have documented this phenomenon (41,42). Earlier research also shows that desexualization is reinforced not only through daily interactions but also through institutional, legal, and policy frameworks. Such systems tend to infantilize women with disabilities, thereby lessening their autonomy by negating their adult status with reference to sexual and social domains (15,18,33,43).

So, where would that leave our findings? The findings suggest that although there is increasing acknowledgment of sexual desire, and challenges of women with disabilities,, this awareness may not be translated into real structural changes. Legislation and changes in public attitudes are often superficial and fail to address the deeper systems that continue to marginalize individuals with disabilities (44).

To move beyond symbolic gestures, there must be actual structural changes behind the shift in discourse. It must translates into rights-based policies, inclusive services, and legal guarantees that in a very active way affirm the sexual autonomy and embodied agency of women with disabilities. If these reforms are not put into place, this gap between public perception and lived experience will remain.

### Community perceptions of sexual organs of women with disabilities

Our study established that there was a widespread acknowledgement among most respondents that women with disabilities had ‘normal’ sexual organs, thus general acknowledgment of their sexual embodiment. Religion was; however, the only factor significantly associated with the perception of the anatomy of women with disabilities.

The research findings, however, contrast with prior research which records women with disabilities as “broken” or deprived of basic physical autonomy (43,45). These dehumanizing descriptions reinforce the perception of women with disabilities as an inherent “other,” abandoned and less than fully human.

Religious faith will often come from local cosmologies, doctrinal teaching, and dominant moral codes. These systems, therefore, play a strong role in determining how non-normative bodies are conceived and regarded across cultural contexts (46).

In many parts of sub-Saharan Africa, the foundations of culture and religion often stand side by side; hence, both cultures and religion establish the notions of what a disability means and its lived-experience.

Building further on this, an increasing number of studies show spiritual interpretations, especially under Christian or Indigenous belief systems, marginalizing women and girls with disabilities (47). These frameworks frequently promote ideals centered on bodily purity, physical perfection, and a divinely ordained order. Such ideals can implicitly or explicitly exclude individuals with disabilities from full moral and social inclusion within their communities (48).

Disability is often symbolically interpreted in African Traditional Religion, such as in ancestral curses, witchcraft, or spiritual imbalance. These beliefs generally invite stigma, social exclusion, and myriad forms of discrimination. Women with disabilities are disproportionately affected by these beliefs, frequently being perceived as transgressing cultural expectations related to fertility, purity, and lineage (49,50).

But care must be taken to avoid generalizing, for Indigenous traditions are not a homogeneity. They also have strong communal care ethics and a collective sense of obligation. However, when these are invoked positively and actively, especially under the auspices of Ubuntu’s foundational philosophy, these traditions can be powerful instruments for advancing disability inclusion and rights (51,52). This internal duality points to the complex and at times, paradoxical nature of Indigenous religious systems, which can easily marginalize within some settings but offer routes to empowerment in others, contingent upon the interpretations and practices embraced in response to their moral frameworks.

### Community perceptions of sexual activity of women with disabilities

The findings of this study indicate that nearly all respondents perceive women with disabilities as sexually active. This normative recognition of sexual activity cuts across demographic variables such as sex, age, religion, education, and prior contact with disability. It suggests the presence of a broadly held community belief in the sexual agency of women with disabilities in Kibra.

This recognition stands in stark contrast to studies that document the characterization of women with disabilities as being uninterested in sexual matters, asexual, or incapable of having sex (53). Instead, the current findings agree with a growing number of studies establishing that women with disabilities are sexually active, just like other women in both low- and high-income settings (12,54,55)

A scoping review of sub-Saharan Africa observed that the majority of prior studies have emphasized the use of contraceptives by women with disabilities, but little on their desire for and engagement in sexual activities (56). However, recent research shows some significant shift. A recent study from Nigeria observes that women with disabilities pursue sexual relationships and negotiate sexual agency willingly in the face of prevalent social stigma and limited access to reproductive health care (57).

Evidence from research in the developed world also validates the occurrence of sex among women with disabilities. Using large-scale U.S. survey data, women of childbearing age with cognitive or multiple disabilities were found to have more than twice the prevalence and significantly higher odds of sexually transmitted diseases compared to women without disabilities, an objective measure of sexual behaviour (54)

In Canada, qualitative adult interviews present rich first-person narratives of people finding and maintaining intimate relationships in the face of systemic obstacles (55). A review of sexual health literature on young adults and adolescents with intellectual disabilities in high-income contexts similarly recognizes “sexual activity” as an emerging theme, only mapping its presence and its articulation of sexual agency (58). Empirical studies attest that although they are under constant stigma and social exclusion, women with disability categorically establish their sexual agency: they pursue intimate relationships, exert sexual demands, and actively take part in sex.

### Community Attitudes towards women with disabilities

The study investigated the extent to which gender, age, religion, education, and type of contact with persons with disabilities shape attitudes toward women with disabilities in Kibra. Some demographic variables exhibited no influence, while others have shown intricate and context-specific interactions.

In terms of **sex**, females in Kibra exhibited slightly higher ATDP scores than did males, but this difference failed to reach significance. These results are contrary to those reported in Nigeria, where being female predicted more positive attitudes (59). Our findings concur with studies from Pakistan (60), Indonesia (61), Ethiopia (62), Greece (63), and Saudi Arabia (64), which also found no statistically significant association between sex and attitude. The variance in findings across cultural settings may be attributed to the fact that sex by itself does not determine a given attitude. Instead, the effect of sex is mediated by social norms and responsibilities; attitudes may be more positive where females are involved extensively in caregiving. Yet, widespread stigma and limited resources can weaken this effect across the broader population.

The ATDP scores by **age** showed no statistically significant difference in our study. This lack of statistical association means that males and females and young adults and older residents have similar attitudes toward women with disabilities. Such uniformity contrasts with findings elsewhere. Age-related variation has been reported in Pakistan and Ethiopia (60,62), and more negative views among older health professionals were found in Saudi Arabia (64). Shorter academic tenure, indicating younger age, has also been linked with more favourable attitudes in Nigeria (59). In our location of study, here in Kibra, there is the possibility that intergenerational interactions shared by the challenges of informal settlement life may provide a unifying factor that cuts across age ranges, occasioning similar attitudes among different age groups.

**Religion** was a statistically significant negative predictor of attitudes toward women with disabilities. Respondents belonging to African Traditional Religion had the lowest mean ranks, much lower than those of any other religious groups. Contrary to our study, no significant effect of religion was found in Pakistan, showing the context-specific nature of religious influence (60). The findings suggest that disability attitudes rely not only on religious identity or lack thereof but also on individual disposition. Both the interpretation and practice of religious belief tend to encourage inclusive or exclusionary disability attitudes.

In our study, another negative predictor of attitudes was **education**, with increasingly positive perceptions of disability with decreasing educational attainment. The results concur with a broad review suggesting that formal education, in and of itself, often may not transform attitudinal dispositions toward the disability issue (65). Contrary to this, Greek nursing students showed greater positive attitudes in advanced stages as disability competencies were gradually integrated into their curriculum (63). On the whole, it appears that attitude formation is more affected by educational content and its pedagogic framing than by the level of education itself.

The strongest and most positive predictor of attitude was considered contact with individuals with disabilities. Caregivers who share daily, affective relationships with people with disabilities have the highest mean rank. The implication is that greater direct interaction leads to a more positive attitude. Our findings align with a systematic review, which observed that contact frequency, intimacy, and emotional reciprocity usually take precedence over mere passive exposure (65). Moreover, having a close friend with a disability inclined one toward holding inclusive attitudes (61), while similar results were observed among colleagues in care settings (62). Personal relationships also break stereotypes more effectively than awareness campaigns (59). In Kibra, where caregiving is done through extended families and community ties, close relationships seem to bring about real change in attitudes. With that backdrop, the studies share the view that building disability-friendly attitudes requires personal, sustained, emotionally meaningful contact with individuals with disabilities rather than broad exposure.

## Conclusion and Contribution

The study sought to evaluate community attitudes and opinions about sexuality among women living with disabilities in the Kibera area of Nairobi. There are generally positive attitudes in the community towards women with disabilities when it comes to their sexual feelings, organs, and activity.

The findings add to disability studies by identifying the complex social and cultural factors, which are context-specific and situational, influencing perception with regard to women’s rights with disabilities in sexual and reproductive matters in informal urban settlements. The continued existence of stigmatizing religious and cultural narratives also indicates a perennial hindrance to the full realization of these very rights.

Limitations to this study include a cross-sectional design (cannot show causality), the dependence on self-reported data, the fact that it was carried out in only one area, which somewhat affects the generalizability of findings and challenges associated with social desirability. Future research may look into how recognition of women with disabilities’ sexuality translates into acceptance and endorsement of their sexual agency and motherhood, preferably through longitudinal and qualitative approaches. In addition, an examination of intervention strategies aimed at tackling cultural and religious stigma could help to craft more effective policies and advocacy programs.

## Data Availability Statement

De-identified data supporting the findings of this study are available upon reasonable request from researchers who meet the criteria for access to confidential data, in accordance with the Kenya Data Protection Act. Because the study includes sensitive information on women’s sexuality in informal settlements, the full data set is not publicly available in order to protect participant privacy. Requests for access should be submitted to the Institutional Scientific and Ethics Review Committee at Tangaza University

